# EMBRACE: Explainable Multitask Burnout Prediction for Resident Physicians using Adaptive Deep Learning

**DOI:** 10.1101/2023.06.24.23291864

**Authors:** Saima Alam, Mohammad Arif Ul Alam

**Affiliations:** Berkshire Medical Center, University of Massachusetts Chan Medical School; University of Massachusetts Lowell, University of Massachusetts Chan Medical School

**Keywords:** resident burnout, mutlitask learning, explainable artificial intelligence

## Abstract

Medical residency is associated with long working hours, demanding schedules, and high stress levels, which can lead to burnout among resident physicians. Although wearable and machine learning-based interventions can be useful in predicting potential burnout, existing models fail to clinically explain their predictions, thereby undermining the trustworthiness of the research findings and rendering the intervention apparently useless to residents. This paper develops, EMBRACE, <u>E</u>xplainable <u>M</u>ultitask <u>B</u>urnout p<u>R</u>ediction using <u>A</u>daptiv<u>E</u> deep learning, that employs a novel framework for predicting burnout that is clinically explainable. At first, we develop, a wearable sensor based improved workplace activity and stress detection algorithm, using deep multi-task learning. Next, we present a novel Adaptive Multi-Task Learning (MTL) framework built on top of our activity and stress detection algorithm, to automatically detect burnout. Additionally, this model also completes the resident burnout survey automatically such a way that it can clinically estimate the same burnout level i.e., clinically explainable and trustworthy estimation. We evaluated the efficacy and explainability of EMBRACE using a real-time data collected from 28 resident physicians (2-7 days each) with appropriate IRB approval (IRB# 2021-017).

## 1. Introduction

Workplace stress is a pervasive issue that affects individuals across various professions and industries. It encompasses the psychological, emotional, and physical strain experienced by employees due to demanding work conditions, excessive workload, and challenging interpersonal dynamics. Recent statistics highlight the magnitude of the workplace stress problem, with studies indicating that a 80% of employees reported feeling stressed at work sometimes and 60% of absenteeism was associated with stress in some ways in that survey [30]. This alarming trend raises concerns about the impact of workplace stress on individuals’ well-being, job satisfaction, and overall quality of life.

Recognizing the detrimental effects of workplace stress, researchers and clinicians have developed clinically validated tools to assess and detect stress levels in workers [31]. These tools typically involve questionnaires and surveys that measure various dimensions of stress, including task load, mental effort, emotion, and perceived stress [32]. While these tools provide valuable insights, they are often limited by their reliance on self-reporting and retrospective assessments, which can be subject to recall biases and may not capture real-time stress experiences [33]. To address these limitations and provide real-time monitoring of workplace stress, wearables and machine learning techniques have emerged as promising solutions. Wearable devices equipped with sensors can collect physiological and behavioral data from individuals throughout their workday, offering continuous and objective measurements of stress-related indicators such as heart rate variability, skin conductance, and physical activity. Machine learning algorithms can then analyze this data and predict stress levels in real-time [5].

Medical residency is undeniably one of the most challenging and demanding workplace stress situations that individuals can experience. Medical residency is a highly challenging and demanding period characterized by extended working hours and schedules [17]. The demanding work schedules and long hours of residency, coupled with work-home interference, create a highly stressful environment that predisposes residents to burnout due to several stressors, including sleep deprivation, conflicts with coworkers, difficulty adapting to a new environment, heavy patient responsibilities, lack of control over schedules, as well as personal traits such as neuroticism or introversion that increase the risk of burnout [1]. Burnout can cause physical symptoms (headache, fatigue, gastrointestinal distress, flu, and sleep and appetite changes) and psychological symptoms (irritability, reduced concentration), as well as behaviors like procrastination, daydreaming, and substance use [2]. Additionally, it can lead to an increased risk of depression, suicidal thoughts, and cardiovascular problems [3]. Moreover, the COVID-19 pandemic has exacerbated the long-standing issue of resident burnout in the United States healthcare system, highlighting the urgent need for interventions to support and protect the well-being of these essential frontline workers before it is too late [4]. The combined use of advanced wearable sensor technologies and Machine Learning (ML) algorithms can facilitate the early identification of burnout, thereby providing an opportunity to prevent its occurrence [5]. Despite their potential benefits, wearable sensors and machine learning-based predictions may suffer from a lack of clinical explainability, potentially leading to mistrust among clinicians and limiting their practical use in real-time clinical settings [7].

This paper introduces a novel framework, *EMBRACE*, for enhancing the prediction and explanation of burnout in residents by utilizing a clinically validated survey that is easily comprehensible and reliable for clinicians. More specifically, our key contributions are:

- In *EMBRACE*, we develop a wearable sensor based improved Workplace Activities (WPAs) and stress recognition framework using deep multitask learning technique. Then, utilizing that, we develop a novel explainable Multitask Learning (MTL) framework to automatically predict burnout and explain the prediction by filling out a clinically validated and trustworthy burnout detection survey tool.
- We validated the accuracy and explainability of our proposed *EMBRACE* framework using a real-time collected data from 28 internal medicine residents (2-7 days each) in a natural hospital duty settings with appropriate IRB approval (IRB# 2021-017).

## 2. Related Works

The use of machine learning (ML) techniques in detecting burnout among resident physicians is a relatively new area of research. While Ecological Momentary Assessment (EMA) has shown effectiveness in predicting burnout among residents [8], incorporating ML methods has the potential to enhance prediction performance [9]. However, real-time burnout prediction necessitates continuous monitoring of health vitals and ML techniques [10–12]. Recent systematic reviews [11, 12] indicate that existing just-in-time burnout prediction techniques utilize biomarkers such as skin temperature, motion-based activities (accelerometers), electrodermal fluctuations, and wristband-based blood volume pulse. Various ML algorithms such as MLP, RF, KNN, SVM, LR, CNN, FCN, Time-CNN, RESNET MLP, CNN-LSTM, MLP-LSTM, Inceptiontime, and others have been employed in these studies [11, 12]. However, a common limitation among these works is the lack of clinical explainability, which has not been adequately addressed in this research field [7, 11, 12].

Many researchers proposed different Interpretable/Explainable AI (XAI) algorithms to make complex AI prediction models explainable that include the Additive Feature Attribution method and the local interpretable model-agnostic explanations (LIME) approach [18]. The shapley additive explanations (SHAP) approach combines LIME with Shapely values to provide explanations for black-box models [14]. Other methods include class activation mapping (CAM) [19], DeepLIFT [20], and layer-wise relevance propagation (LRP) [21] for interpreting convolutional neural networks (CNNs). In healthcare, explainable AI applications have been developed for interpreting imaging studies and real-time predictions [22]. One previous work proposed interpretable ML techniques for stress prediction using wearables, but it only provided a simplistic representation of top features based on SHapley Additive exPlanations (SHAP), which lacks clinical significance [13]. Adapa et. al., proposed [15] a supervised machine learning method to predict burnout among resident physicians that takes a bunch of surveys to understand different workplace problems and activities related, and, based on that longitudinal surveys on personal, physical, workplace environmental and physiological status measures, and performed supervised machine learning approach to identify some highly correlated factors (emotional exhaustion, depersonalization, race demographics etc.). *EMBRACE* offers both efficient burnout prediction as well as clinically validated survey filling out method hypothesizing that clinical survey of burnout estimation is explainable and trustworthy among resident physicians.

## 3. EMBRACE Framework

EMBRACE framework consists of two core components: algorithm for detecting workplace activity and stress; and, adaptive algorithm for detecting burnout level and explanation. Fig. 1 presents a schematic diagram of our proposed framework.

**Figure 1.**
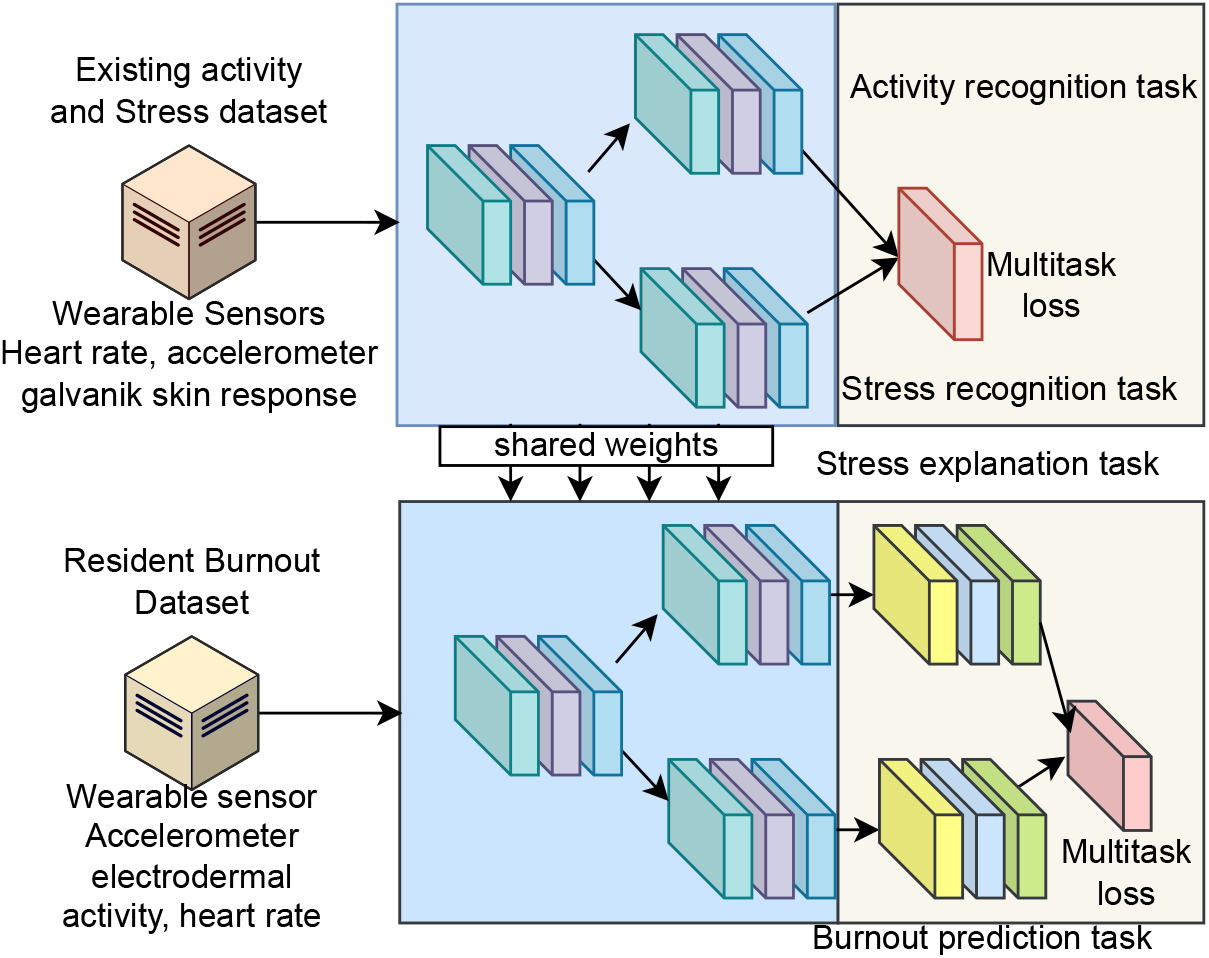
The schematic diagram of proposed framework.

### 3.1. Detecting Workplace Activity and Stress

A multitask deep learning framework for wearable sensor-based activity and stress detection involves training a single model to simultaneously perform multiple tasks, specifically activity recognition and stress level classification. The framework combines both tasks into a single neural network architecture, allowing shared representations to be learned and leveraging the complementary information present in the data.

#### 3.1.1. Input Data

The input data consists of time-series sensor readings from wearable devices, denoted as *X* ∈ ℝ^*T* × *N*^, where *T* represents the length of the time series and *N* is the number of sensor channels.

#### 3.1.2. Activity Recognition Task

Activity recognition aims to predict the activity type based on sensor data. The predicted activity labels are denoted as 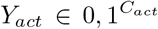, where *Cact* represents the number of activity classes. The output layer for activity recognition is defined as:

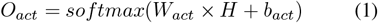

where *H* represents the shared hidden representations obtained from the network, *Wact* is the weight matrix, and *bact* is the bias term specific to the activity recognition task.

#### 3.1.3. Stress Level Classification Task

Stress level classification aims to predict the stress level based on sensor data. The predicted stress labels are denoted as 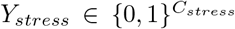, where 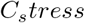 represents the number of stress level classes. The output layer for stress level classification is defined as:

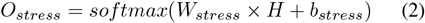

where *H* represents the shared hidden representations obtained from the network, *W*_*stress*_ is the weight matrix, and *b*_*stress*_ is the bias term specific to the stress level classification task.

#### 3.1.4. Shared Representation Learning

The shared representation learning module learns a representation that captures both activity and stress-related patterns in the input data. This module consists of a combination of one convolutional neural networks (CNN) with 32 hidden nodes each and two Long Short Term Memory (LSTM) layers with 64 hidden nodes each, to extract meaningful features from the input time series. The final fused hidden representation obtained from this module is denoted as *H*.

#### 3.1.5. Loss Function

The multitask loss function combines the losses from both tasks to jointly optimize the model. The loss function is defined as a combination of activity recognition loss (*L*_*act*_) and stress level classification loss (*L*_*stress*_), weighted by respective task-specific coefficients (*α* and *β*):

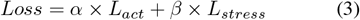

#### 3.1.6. Learning

The model is trained using backpropagation and gradient descent optimization techniques, minimizing the multitask loss function. The shared representation learning module and task-specific layers are updated jointly during training. By training the multitask deep learning framework, the model learns to extract relevant features from the wearable sensor data and simultaneously perform activity recognition and stress level classification tasks. This joint learning approach enables the model to leverage the shared representations and potentially improve the performance of both tasks compared to training separate models.

### 3.2. Burnout Prediction and Explanation

To build a multitask few-shot deep domain adaptation framework based on the previous framework, we will adapt it to the scenario where wearable sensor data serves as input, the source domain involves multitask stress and activity recognition, and the target domain focuses on predicting the answers to a multitask Mini-Z survey questionnaire [23] and burnout prediction. The objective is to estimate the overall burnout scale class based on the Mini-Z survey questions’ answers. We describe this model as follows:

#### 3.2.1. Preliminaries

In this framework, we have similar input data representation where the source domain framework is the previously described multitask deep learning architecture for stress and activity recognition tasks. The model architecture includes shared representation learning, output layers for activity recognition (*O*_*act*_) and stress level classification (*O*_*stress*_), and corresponding labels *Y*_*act*_ and *Y*_*stress*_. In the target domain, the focus shifts to predicting the answers to the multitask Mini-Z survey questionnaire. The objective is to estimate the overall burnout scale class based on the answers to the Mini-Z survey questions. For each Mini-Z survey question, a separate output layer is defined in the neural network architecture. The output layer for predicting the answer to question *i* is denoted as *O*_*i*_ = *f* (*W*_*i*_*H* + *b*_*i*_), where *H* represents the shared hidden representations obtained from the network, *W*_*i*_ is the weight matrix specific to question *i, b_i_* is the bias term associated with question *i*, and *f* is an appropriate activation function. The estimated overall burnout scale class is derived from the answers to the Mini-Z survey questions. This has been achieved by defining a range of total Mini-Z survey questions’ answers and mapping it to specific burnout scale classes.

#### 3.2.2. Multitask Adaptive Loss Function

The multitask loss function for the target domain includes the task-specific loss for Mini-Z survey questions prediction (*L*_*miniZ*_) and the overall burnout scale class loss (*Lburnout*), weighted by respective task-specific coefficients (*γ* and *δ*). The loss function is defined as:

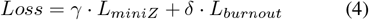

where *L*_*burnout*_ is the cross-entropy loss for the overall burnout scale class estimation and *L*_*miniZ*_ is the *R*^2^ loss metrics. R-squared is a goodness-of-fit measure for regression models. This statistic indicates the percentage of the variance in the dependent variable that the independent variables explain collectively. R-squared measures the strength of the relationship between your model and the dependent variable on a convenient 0 – 100% scale. *R*^2^ loss can be represented as follows:

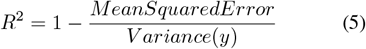

#### 3.2.3. Few-shot Domain Adaptation

Few-shot domain adaptation aims to transfer knowledge from the source domain to the target domain, even when labeled data in the target domain is limited. We modify Model-Agnostic Meta-Learning (MAML) algorithm [24] according to our multitask source and target problem, which allows the model to quickly adapt to new tasks using a few labeled samples. The modified MAML algorithm includes initialization of model parameters and source domain training. Then, the few-shot domain adaptation includes selecting few target samples with labels to define new target task with cloned source model’s parameters. Then, for each target domain task, we perform a few gradient update steps on target parameters using few samples and compute target task specific loss in the inner loop; and compute gradient of task-specific target loss with respect to source parameters and update it. Finally, we evaluate the adapted target task model using Mini-Z survey answers-based prediction 1.

##### Algorithm 1 Multitask Few-Shot Deep Domain Adaptation with Model-Agnostic Meta-Learning (MAML)

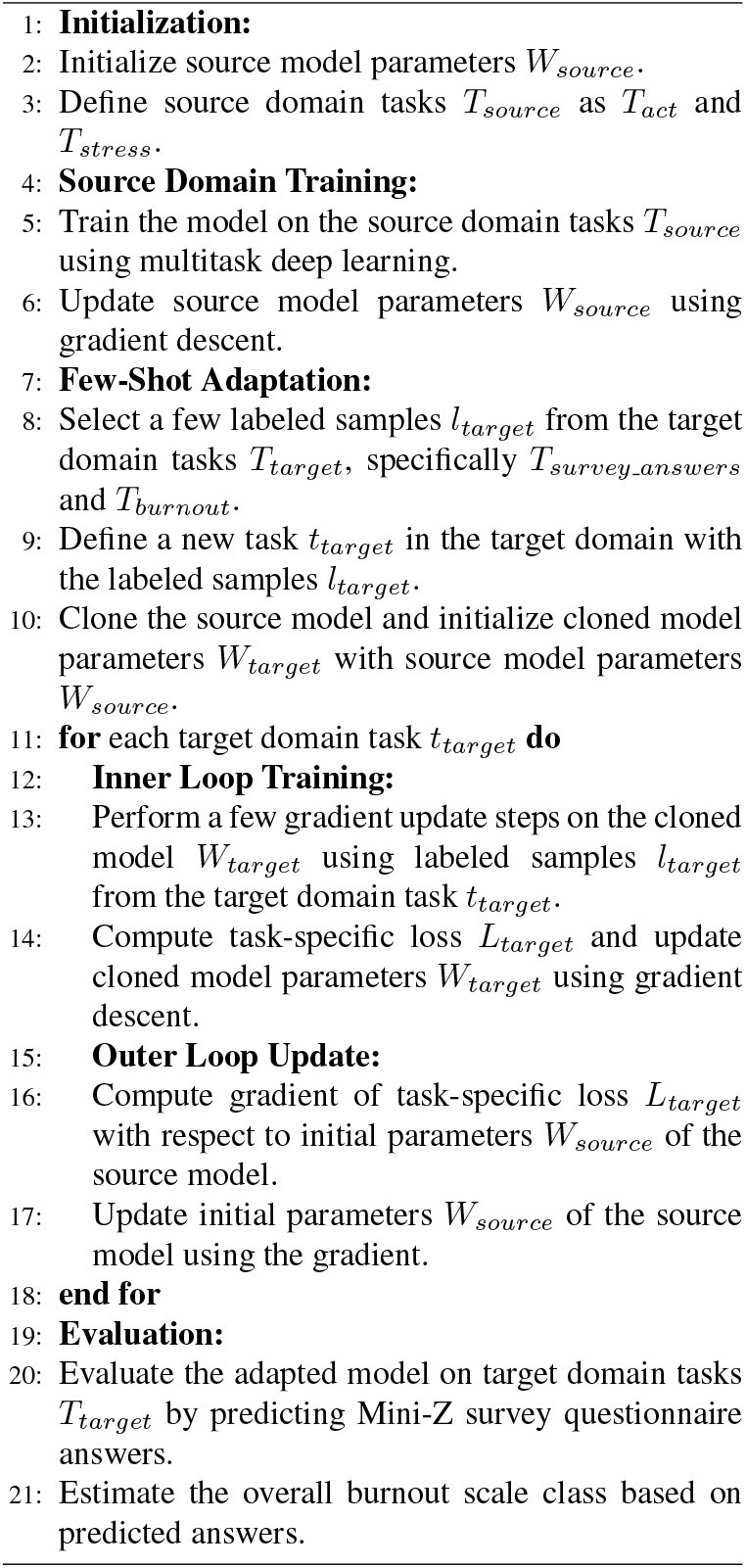

## 4. Experimental Evaluation

This section describes our experimental evaluation on *EMBRACE* framework.

### 4.1. Datasets

We have utilized one existing datasets as source for developing our stress and activity recognition and performance, and collected our own dataset for developing Multitask Few Shot Deep Domain Adaptation with MAML model.

#### D1 (SWELL-KW Dataset)

We utilized SWELL-KW dataset [25] as source that contains sensor data (accelerometer, heart rate, galvanik skin response), activity labels and four clinical tool estimated stress assessment ground truth. The dataset was collect from 25 people (3 hours each) performed workplace activities under manual stressors such as email interruptions and time pressure. The data has been annotated with activities (via recorded videos), subjective experience on task load, mental effort, emotion and perceived stress was assessed with validated questionnaires (NASA-TLX [26], RSME [27], SAM and PSS [29]) as ground truths.

#### D2 (Burnout Dataset)

With the appropriate Institutional Review Board (IRB) approval (IRB# 2021-017), we recruited 28 internal medicine resident physicians (avg. age 27.5 with std 3.5) from a renown teaching-based medical center from different year (PGY1, PGY2, PGY3). Each participant was asked to wear Empatica E4 watch from the beginning of their daily duty until the end of the day. To assess clinical burnout among resident physicians, we utilized Mini-Z Burnout Survey [23]. The Mini-Z (2.0) burnout assessment comprises a set of 10 questions that utilize 5-point Likert scales, along with an additional open-ended question. These items aim to evaluate three key outcomes, namely burnout, stress, and satisfaction, while also exploring seven factors that contribute to burnout. These factors encompass work control, work chaos, teamwork, alignment of values with leadership, documentation time pressure, EMR (Electronic Medical Record) use pressure, and EMR proficiency. There are three different burnout scales are estimated from these 10 answers:

1. Joyful measure: Add all points from the 10 items for a total score, range 10–40 points. A score ≥20 is considered representative of a joyful work environment
2. Satisfaction scale: Add all points from Q1, Q2, Q3, Q4, range 4–25 points. A score ≥20 is considered a highly supportive environment.
3. Stress scale: Add all points from Q5, Q6, Q7, Q8, range 4–25 points. A score ≥20 is considered a low stress environment with reasonable EMR pressures.

A text notification of online MiniZ burnout survey [23] form was sent to their cell phone at 7PM everyday that must be submitted by midnight. The Empatica E4 data as well as submitted MiniZ burnout survey data are stored in a secured HIPAA compliant server with proper de-identification. We performed this Mini-Z survey evaluation before the collection had started to calculate participants’ baseline burnout profile.

### 4.2. Tasks Definitions

There are two tasks involved in the source dataset (D1), Task 1 (*T*_*act*_): 5-class activity recognition (writing reports, making presentations, reading e-mail, searching for information and others) and Task 2 (*T*_*stress*_): 3-class stress level recognition (neutral, interruption and Time pressure). On the other hand, there are four tasks involved in the target dataset, D2, Task 1 (*T*_*survey_answers*_): 10-class regression problem to fill-out survey questions and, Task 2 (*T*_*burnout*1_): 2-class overall measure (joyful work environment or not), Task 3 (*T*_*burnout*2_): 2-class satisfaction scale (highly supportive work environment or not), and, Task 4 (*T*_*burnout*3_): stress scale (low stress environment with reasonable EMR pressure or not). In Fig. 1, we presents three burnout tasks (Task 2, Task 3 and Task 4) into a single domain.

#### Implementation

Our proposed model was implemented using Python’s Keras library with the TensorFlow backend. For the regression task, denoted as 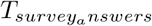, we employed the RMSE loss function. In contrast, for the classification tasks, which encompassed the remaining tasks, we utilized categorical cross-entropy loss. These loss functions were employed while jointly training the few-shot MAML algorithm.

The optimization of our system was performed using the Adam optimization function with a learning rate of 1 × 10^*−*3^. The selection of the optimized learning rate and the weighting parameter *β* (set to 0.25) was achieved through hyperparameter tuning. The learning model of our framework was executed on a server equipped with a cluster of three Nvidia GTX GeForce Titan X GPUs and an Intel Xeon CPU (2.00GHz) processor, along with 12 gigabytes of RAM.

For training the multitask stress and workplace activity recognition framework, we utilized the D1 dataset (SWELL-KW Dataset) as input. This dataset included readings from wearable sensors such as accelerometers, heart rate monitors, and galvanic skin response sensors. The framework was trained to address two tasks. To adapt the shared module of the target adaptive multitask explainable burnout prediction, we employed the trained weights for initialization (domain adaptation). Subsequently, we replaced the inputs in our collected dataset, D2, with readings from wearable sensors such as accelerometers, heart rate monitors, and electrodermal activity sensors. Additionally, we modified the output layer to accommodate the four aforementioned task problems.

### 4.4. Prediction Performance Evaluation

The conventional 10-fold cross validation approach is not suitable for sequential data. Therefore, to train and assess the performance of our proposed *EMBRACE* framework, we adopt a different approach. We partition the entire sequential dataset into two halves. Subsequently, we randomly select a sequence of data from the first half as the training sample, and another random sequence from the second half as the testing sample. This process is repeated ten times to generate ten distinct pairs of training and testing data sequences. To evaluate the performance of our algorithm, we utilize balanced accuracy, precision, recall, and F1 measure as metrics. These metrics provide a comprehensive assessment of the algorithm’s effectiveness. Additionally, we calculate the standard deviation of all these metrics to evaluate the presence of overfitting.

Table 1 and Table 2 present detailed performance results of the multitask activity and stress detection algorithm implemented in the *EMBRACE* framework on the source dataset, D1 (SWELL-KW dataset).

**Table 1.** Workplace activities (writing reports, making presentations, reading e-mail, searching for information and other random works) accuracy performance of source dataset, D1, on EMBRACE framework.

| Activities | Accuracy | Precision | Recall | F1 measure |
| --- | --- | --- | --- | --- |
| Writing | 97.5±.02 | 95.1±.02 | 97.5±.02 | 97.8±.01 |
| Present | 92.4±.1 | 92.6±.15 | 92.4±.1 | 93.7±.1 |
| E-mail | 85.3±1.1 | 86.3±.9 | 85.3±1.1 | 87.8±.8 |
| Searching | 89.4±1.2 | 90.8±0.6 | 89.4±1.2 | 90.3±1.3 |
| Others | 85.3±1.1 | 87.8±1.4 | 85.3±1.1 | 88.1±1.5 |
| Overall | 91.6±0.9 | 93.1±0.5 | 91.6±0.9 | 93.9±0.2 |

**Table 2.** Stress level (neutral, interruptions and time-pressure) accuracy performance of source dataset, D1, on EMBRACE framework.

| Stress Levels | Accuracy | Precision | Recall | F1 measure |
| --- | --- | --- | --- | --- |
| Neutral | 99.5±.00 | 98.2±.01 | 99.5±.00 | 99.1±.01 |
| Interrupt | 94.1±.07 | 95.4±.01 | 94.1±.07 | 96.3±.06 |
| Time | 91.2±.09 | 92.7±.09 | 91.2±.09 | 92.8±.08 |
| Overall | 94.7±.02 | 94.7±.02 | 94.7±.02 | 95.1±.01 |

Table 1 displays the overall accuracy, precision, recall, and F-1 measure for workplace activity recognition, which achieve values of 91%, 93%, 91%, and 93% respectively. These results are accompanied by reasonably low standard deviations, indicating no evidence of overfitting. Notably, the classification of writing reports achieves a significantly higher accuracy of 97% compared to other tasks.

Table 2 reports the overall accuracy, precision, recall, and F-1 measure for stress level recognition, with values of 94%, 94%, 94%, and 95% respectively. Similar to the activity recognition results, the standard deviations are reasonably low, suggesting no overfitting. It is worth highlighting that the classification of neutral stress levels demonstrates an impressive accuracy of 99%, outperforming the other stress level classifications.

Table 3 and Table 4 present the R-squared measure for survey questionnaire completion and the burnout prediction accuracy performance of the adaptive multitask burnout prediction and explanation framework, referred to as *EMBRACE*.

**Table 3.** Mini-Z survey questionnaire specific answer regression mean squared error (MSE) on our collected dataset, D2, using adaptive multitask learning part of EMBRACE framework.

| Questions | Accuracy | Precision | Recall | F1 measure |
| --- | --- | --- | --- | --- |
| Q1 | 78.5±.9 | 79.5±.9 | 78.9±.8 | 80.6±.9 |
| Q2 | 75.8±.7 | 77.4±.8 | 75.3±.9 | 76.4±.9 |
| Q3 | 69.5±1.9 | 70.6±1.1 | 70.5±1.2 | 71.6±1.0 |
| Q4 | 84.6±.9 | 87.8±0.7 | 84.6±0.9 | 86.5±0.9 |
| Q5 | 97.5±.01 | 98.2±.01 | 97.5±.01 | 98.3±.01 |
| Q6 | 96.3±.01 | 95.9±.02 | 96.3±.01 | 97.1±.02 |
| Q7 | 93.6±.02 | 94.8±.03 | 93.6±.02 | 93.6±.01 |
| Q8 | 90.5±.3 | 88.5±1.1 | 90.4±.2 | 91.3±.8 |
| Q9 | 86.5±.9 | 87.1±1.2 | 85.9±.5 | 88.8±.9 |
| Q10 | 90.2±1.0 | 89.4±1.1 | 90.2±1.0 | 91.5±.8 |
| Overall | 87.7±0.5 | 88.3±0.8 | 87.6±0.4 | 88.8±0.7 |

**Table 4.**
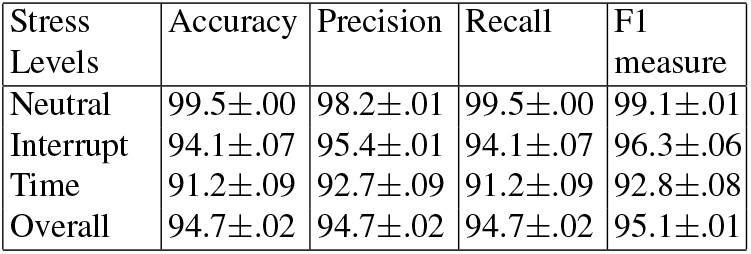
Three kind of burnout measures (Joyful measure, Satisfaction scale and Stress scale), D2, using adaptive multitask learning part of EMBRACE framework.

The results demonstrate significant overall accuracy, precision, recall, and F-1 measures of 87%, 88%, 87%, and 88% respectively. Although certain questionnaires (Q1, Q2, and Q3) show exceptionally poor R-squared measures, the final accuracy of stress level prediction (as shown in Table 4) is not significantly affected due to the efficient design of our adaptive multitask learning framework.

The overall accuracy, precision, recall, and F-1 measures for stress level prediction are 94%, 94%, 94%, and 95% respectively. Furthermore, both Table 3 and Table 4 reveal reasonably low standard deviations, indicating a lack of strong evidence for overfitting in our models.

### 4.5. Discussion

To enhance the clarity of our findings, we applied the multitask workplace activity and stress detection algorithm to our collected dataset, referred to as D2. In order to gain deeper insights, we conducted correlation analysis among various predicted variables, including workplace activities, stress levels, the Mini-Z questionnaire responses, and burnout measures. The correlation analysis results are depicted in Figure 2, where darker colors indicate lower correlation values.

**Figure 2.**
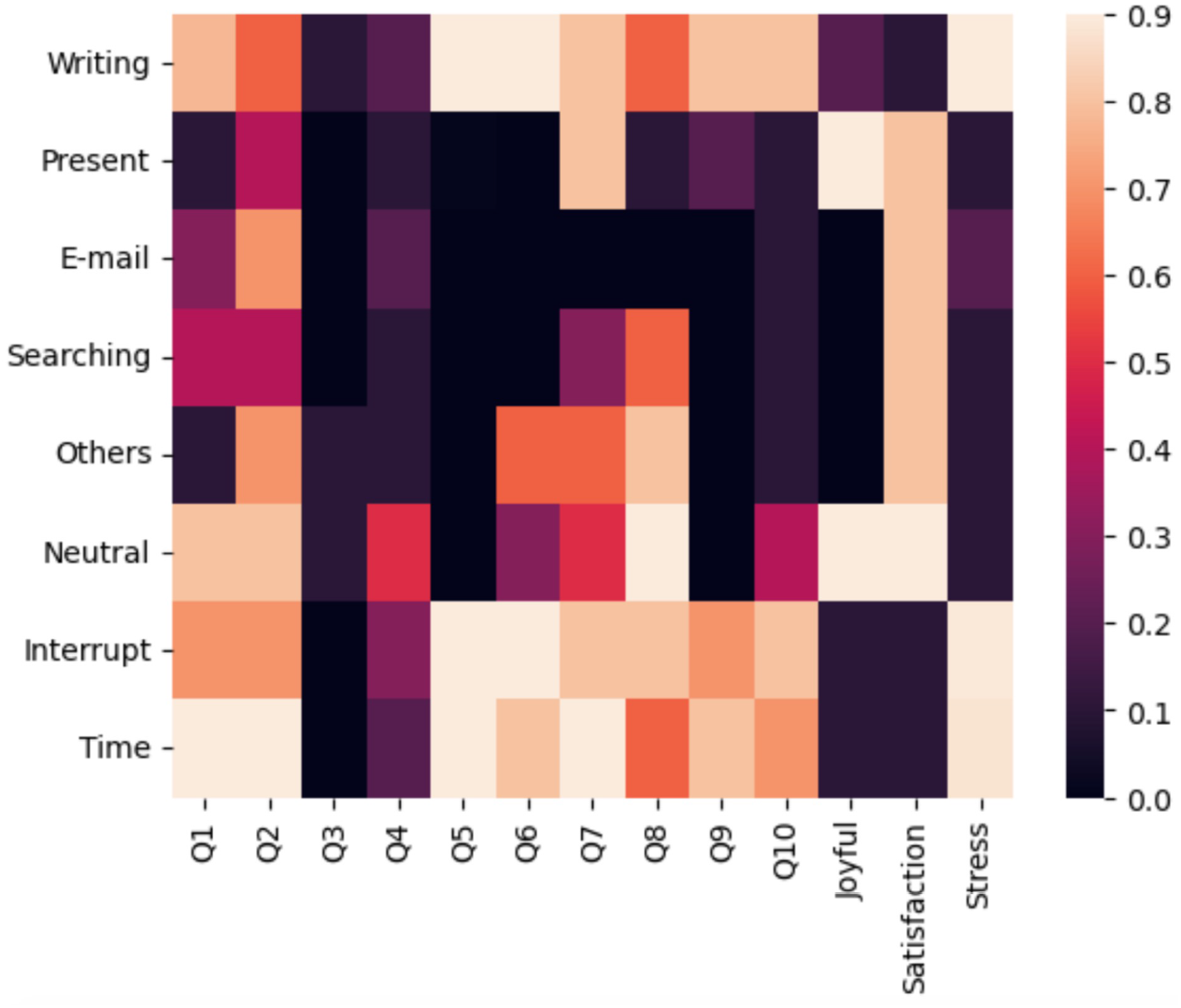
Correlation heatmap among detected workplace activities and stress; with, Mini-Z survey questionnaires and estimated bunout measures.

The questionnaires used in the analysis include the following items: Q1: Overall, I am satisfied with my current job; Q2: Using your own definition of “burnout,” please circle one of the answers below; Q3: My professional values are well aligned with those of my department leaders; Q4: The degree to which my care team works efficiently together; Q5: I feel a great deal of stress because of my job; Q6: The amount of time I spend on the electronic medical record (EMR) at home; Q7: Sufficiency of time for documentation is; Q8: Which number best describes the atmosphere in your primary work area?; Q9: My control over my workload is; and Q10: The EMR adds to the frustration of my day. Our analysis reveals several notable findings. Firstly, it is evident that highly interruptive and time-pressured work activities exhibit strong correlations with most of the questionnaires and stress levels. This suggests that an increase in interruptions and time pressures in the workplace is associated with higher levels of stress and negative responses on the questionnaires. However, we also observed a positive correlation between a neutral (stress-free) state and job satisfaction, as well as a pleasant workplace environment.

Furthermore, when examining specific workplace activities, we found that writing notes demonstrated a strong correlation with stress levels. This indicates that an increase in the frequency of writing notes during work is associated with higher levels of stress among residents. Additionally, writing notes showed high correlations with questionnaire items Q5 and Q6, both of which are related to documentation and electronic medical record (EMR) writing.

Lastly, it is worth noting that presentation activities in the workplace exhibited a high correlation with a joyful work environment, suggesting that engaging in presentations contributes to a positive and pleasant atmosphere.

## 5. Conclusion and Future Work

This paper introduces a novel approach to estimate and explain burnout measures using adaptive multitask learning. The estimation is conducted by administering burnout surveys, and the results are further explained through correlation analysis between our proposed workplace activity recognition and stress measures, along with our predictive model parameters. Although our framework achieves notable accuracy, we were unable to compare it with existing frameworks as there were no similar frameworks available in the current state-of-the-art literature. Furthermore, we did not conduct a satisfaction study on explainability, which would require longitudinal data from a significantly larger number of participants. Our framework is the first of its kind to offer an explanation of wearable-based burnout measures, thus introducing a new dimension in the field of explainable machine learning.

## Data Availability

All data produced in the present study are available with appropriate deidentification upon reasonable request to the authors

